# Definition of sinonasal and otologic exacerbation in patients with primary ciliary dyskinesia - an expert consensus

**DOI:** 10.1101/2024.03.08.24303910

**Authors:** Myrofora Goutaki, Yin Ting Lam, Andreas Anagiotos, Miguel Armengot, Andrea Burgess, Raewyn Campbell, Mathilde Carlier, Nathalie Caversaccio, Neil K. Chadha, Berat Demir, Sinan Ahmed D. Dheyauldeen, Onder Gunaydin, Amanda Harris, Isolde Hayn, Deniz Inal-Ince, Eric Levi, Trini Lopez Fernandez, Jane S. Lucas, Bernard Maitre, Anne-Lise ML Poirrier, Lynne Schofield, Kazuhiko Takeuchi, Christine van Gogh, Nikolaus E. Wolter, Jean-François Papon

**Author notes:** Correspondence: Myrofora Goutaki, Institute of Social and Preventive Medicine, University of Bern, Mittelstrasse 43, 3012 Bern, Switzerland.

## Abstract

**Background:** Recurrent infections of the nose, sinuses, and ears are common problems for people with primary ciliary dyskinesia (PCD). While pulmonary exacerbations in PCD are defined, there is no definition for Ear-Nose-Throat (ENT) exacerbations, a potential outcome for research and clinical trials.

**Methods:** We set up an expert panel of 24 ENT specialists, respiratory physicians, other healthcare professionals, and patients to develop consensus definitions of sinonasal and otologic exacerbations in children and adults with PCD for research settings. We reviewed the literature and used a modified Delphi approach with four electronic surveys.

**Results:** Both definitions are based on a combination of major and minor criteria, requiring three major or two major and at least two minor criteria each. Major criteria for a sinonasal exacerbation are: 1) reported acute increase in nasal discharge or change in colour; 2) reported acute pain or sensitivity in the sinus regions; 3) mucopurulent discharge on examination. Minor criteria include: reported symptoms; examination signs; doctoŕs decision to treat; improvement after at least 14-days. Major criteria for the otologic exacerbation are: 1) reported acute ear pain or sensitivity, 2) reported acute ear discharge, 3) ear discharge on examination, 4) signs of otitis media in otoscopy. Minor criteria are: reported acute hearing problems; signs of acute complication; doctoŕs decision to treat.

**Conclusion:** These definitions might offer a useful outcome measure for PCD research in different settings. They should be validated in future studies and trials together with other potential outcomes, to assess their usability.

## Introduction

Dysfunction of motile cilia due to genetic mutations leads to a wide range of symptoms including multiple organ systems in patients with primary ciliary dyskinesia (PCD).[1, 2] Despite the clinical heterogeneity, the greatest impact of impaired mucociliary clearance is seen on the respiratory tract and the ears.[3] Patients present with persistent wet cough and recurrent lower airway infections, progressing with time to irreversible lung damage.[3] Inadequate clearance of mucus, pathogens, and debris in the nose and sinuses, as well as in the eustachian tube and middle ear, leads to bacteria growing in the mucus-clogged airways. Consequently, patients experience recurrent episodes of sinonasal infections, and the risk of sinonasal disease increases with age, with chronic rhinosinusitis (CRS) becoming a common feature as disease progresses.[4–6] From the ears, recurrent episodes of acute otitis media (AOM) often progress to severe bilateral otitis media with effusion (OME) and conductive hearing impairment.[7–11] Acute infections of the nose, sinuses, and ears in PCD, usually involve already impaired upper airways, with a more complicated pathophysiology and course compared to common acute upper airway infections.

Respiratory exacerbations are a significant determinant of morbidity and subsequent care requirements of people with chronic respiratory diseases. They are typically characterised by deterioration of the patient’s clinical condition, most often due to viral or bacterial infections or exposure to other triggering factors. Exacerbations often require additional management and have significant effects on disease progression, severity, and patientś quality of life.[12–15] For clinical and epidemiological research, exacerbations are important outcomes, in measuring burden of disease or response to treatments.[16, 17] In PCD, pulmonary exacerbations have been defined, and were recently included in a core outcome set for pulmonary disease interventions,[18, 19] in the framework of the BEAT-PCD clinical research collaboration supported by the European Respiratory Society (ERS).[20, 21] The existing definition excluded upper respiratory tract exacerbations because they often occur independently from lower respiratory tract exacerbations and have different prognosis.[18] Therefore, despite their impact on severity of PCD, there is still no definition for Ear-Nose-Throat (ENT) exacerbations. This lack is an important gap as clinical outcomes capturing ENT disease in PCD are even fewer than for lung disease.[22]

Using an international panel of specialists involved in PCD care, we aimed to develop a consensus definition of ENT exacerbations for children and adults with PCD, participating in clinical research.

## Methods

### Participants and purpose of the consensus

We established an expert panel consisting mainly of ENT specialists, with expertise in managing children and adults with PCD. We invited specialists from PCD reference centres in Europe, North America, Australia, and Japan, and encouraged invited participants to suggest further members ensuring wide international representation (Supplementary Table S1). Additionally, we invited a paediatric and an adult pulmonologist, involved in the consensus group of the pulmonary exacerbations definition,[18] and other healthcare professionals involved in PCD patient care and research. The panel was completed by two patient representatives, an adult with PCD and a parent of a child with PCD; 24 members in total, representing 13 countries. To ensure significant patient involvement and input from the people who experience first-hand these exacerbations, we also set up a parallel group of patient and parent volunteers, with support from the European Lung Foundation (ELF),[23] who did not join the consensus panel, but provided feedback and were encouraged to participate in the surveys. The activities of the panel and the patient group were coordinated by two facilitators, a clinical epidemiologist with expertise in PCD research (MG) and a PCD PhD candidate (YTL); the latter did not participate in voting. An initial virtual panel meeting refined the aims and proposed methodology. It was concluded that standardised definitions for PCD were missing and decided unanimously to produce two separate definitions: one for sinonasal exacerbations and one for otologic exacerbations. Our goal was to establish definitions meant to be used in research and clinical practice.

### Literature search

We conducted a systematic literature search of publications referring to ENT exacerbations in patients with PCD, or separately sinonasal and otologic exacerbations. Given the anticipated limited pre-existing literature on the topic, our search strategy was expanded at the outset to include other areas with common characteristics, in particular CRS. We searched PubMed for studies published between January 2012 and December 2021 using the following keywords: ciliary dyskinesia, primary OR immotile cilia syndrome OR Kartagener/ AND exacerbat* OR infect* OR acute/ AND sinus* for sinonasal exacerbations, or otit* OR ear or otol*, for otologic ones. We simplified the terms, excluding PCD specific keywords, to expand on other diseases excluding the PCD related keywords. We did not exclude any publication type or language.

### Reaching a consensus

A modified Delphi approach with online (eDelphi) surveys was used. Initial literature search results revealed that identified pre-existing definitions did not cover the need for PCD-specific definitions but should be used as a starting point for the first eDelphi survey. Based on these definitions and the panel consensus, we identified important components for definitions of sinonasal and otologic exacerbations. For each survey, participants received detailed instructions and a link via e-mail, then two reminders to respond within two and three weeks. The panel decided that at least an 85% response rate would be required to proceed to the next survey and that 80% of agreement would signify consensus; however, possibility of accepting lower agreement as consensus was left open provided that the panel would be informed and there would be no veto against it. Each survey (eDelphi survey Supplementary material 1-4) included different types of questions to reach consensus initially on the included components for each definition and subsequently on the details of specific components e.g. elements of included components such as specific symptoms or signs. Each survey was designed based on the results of the previous survey and included a summary of these for the paneĺs information. Participants were asked to explain their opinions in free text boxes, particularly where consensus was not achievable despite high agreement, so statements could be clarified and modified in the next round. The number of surveys was not predefined, but ultimately 4 surveys were required. A virtual meeting was organised with MG and the patient group to explain details of the project to the patient or parent members and provide any answers to their questions. Replies remained anonymous to the panel and only the facilitators had access to identifying information. After the eDelphi surveys were completed, the results of the final survey were circulated among the panel to ensure all members agreed with the final definitions.

## Results

### Literature search

Our search resulted in total of 2352 abstracts related to sinonasal and 2208 to otologic exacerbations respectively. No abstracts with definitions specific to PCD were identified. After excluding duplicates and screening the abstracts, we identified 24 manuscripts that referred to sinonasal exacerbations. By searching their references 6 additional manuscripts were identified, 30 in total, including one systematic review.[24] A summary of definitions used in literature for sinonasal exacerbation in patients with CRS in the identified studies is presented in Table 1.[24–53] These definitions were discussed at a virtual expert panel meeting and the elements they used were considered for developing the initial survey. No study fulfilled the criteria of otologic exacerbation of a chronic condition.

**Table 1:** Summary of definitions used in literature for sinonasal exacerbation in patients with chronic rhinosinusitis.

| Definition | References |
| --- | --- |
| Acute increase in **severity** of sinus disease symptoms | Armbruster 2021 |
| Sudden **worsening** of CRS symptoms with a **return** to baseline symptoms, often after **treatment** | Orlandi 2020; Bleier 2021 |
| Acute worsening of pre-existing CRS symptoms with **subsequent return to baseline** symptoms **with or without endoscopic evidence** | Orlandi 2020; Makary 2021 |
| Previous **diagnosis of CRS** exists, and a sudden worsening of symptoms occurs, with a **return to baseline symptoms following treatment** | Orlandi 2016; Philpott 2021; Wu 2019; Yan 2018; Barshak 2017 |
| Presence of **purulence on endoscopy** during a symptomatic exacerbation of CRS | Orlandi 2016; Vandelaar 2019 |
| Sudden **worsening of pre-existing** CRS symptoms is called a CRS exacerbation | Laulajainen-Honigsto 2020 |
| Diagnosis of chronic rhinosinusitis and acute exacerbation of CRS according to the criteria described in the "European Position Paper on Rhinosinusitis and Nasal Polyps (EPOS) | Fokkens 2020; Yaniv 2020 |
| Previous **diagnosis of CRS** but were experiencing **acute** exacerbation **of symptoms** | Fokkens 2012; Ghadersohi 2020; Kuiper 2018 |
| **Acute** worsening of symptoms with return to baseline, often requiring a transient escalation in **treatment**, such as a course of oral antibiotics or corticosteroids | Fokkens 2012; Phillips 2019; Kuiper 2018; Phillips 2018 |
| **Acute** exacerbation of CRS was defined as having received an **antibiotic** prescription **for worsening sinus symptoms** | Kwah 2020 |
| Acute exacerbations among **surgically managed** CRS patients were defined as a post-endoscopic sinus surgery, after **90 days post-op** | Denneny 2018 |
| Acute bacterial CRS exacerbations (**patient-reported** sinus **infections** and CRS-related **antibiotic use**) | Sedaghat 2018 |
| Sudden **worsening of baseline** symptoms (or developing **new symptoms**) in a patient with an established CRS diagnosis | Lopatin 2018 |
| Sudden worsening of the baseline CRS with either **worsening or new** symptoms. Typically, the acute (not chronic) symptoms **resolve completely** between exacerbations | Brook 2016 |
| **Worsening**, with subsequent **resolution**, of symptoms in a patient carrying the **diagnosis of CRS** | Merkley 2015 |
| Defined by minimum **SNOT-20** score of 1.0 on scale of 0 to 5 | Jiang 2015 |
| Worsening of symptoms: facial **pain** or pressure, nasal **obstruction**, nasal **discharge** | Rosenfeld 2015; Beswick 2020 |
| Presence of **increased nasal congestion**, and facial **pain**; **increased sinonasal discharge**; usually presence of an unscheduled sick visit | Zemke 2019, Wu 2020 |
| **Acute** exacerbation of CRS was defined in a patient in whom a previous diagnosis of CRS exists, and a **sudden worsening of symptoms** occurs, with a **return to baseline** symptoms **following treatment** | Orlandi 2016, Wu 2020 |
| Natural exacerbation was defined as **patient-reported worsening of sinonasal symptoms** (i.e. runny nose, nasal congestion, and nasal obstruction) | Divekar 2015, Wu 2020 |
| History of **sudden worsening of preexisting symptoms** suggests an acute exacerbation of CRS, which should be diagnosed by similar criteria and treated in a similar way to acute rhinosinusitis | Fokkens 2012, Wu 2020 |
| Self-reported **medication use** (antibiotics and oral corticosteroids) for **worsened nasal and sinus symptoms**; self-reported **worsened purulence** in the past 4 weeks | Kuiper 2018, Wu 2020 |
| Systemic **antibiotics**; systemic **corticosteroid**; plans for a **semi-urgent surgical intervention**; emergency department or **urgent care visit**, or a **hospitalization** for CRS | Wu 2020 |
| **Worse nasal symptoms** | Wu 2020 |

CRS: chronic rhinosinusitis, SNOT-20: Sino-Nasal Outcome Test

### eDelphi surveys

Response rates to the eDelphi surveys ranged between 88 and 100% (Supplementary Table S1). In addition, two to five members of the patient group completed each survey. In survey 1, the panel assessed opinions about the importance of sinonasal and otologic exacerbations for people with PCD and components that should be included in the exacerbation definitions. Consensus was reached that exacerbations from the nose and sinuses are an important problem for both adults and children with PCD, they impact the quality of life of people with PCD and can be an important outcome measure for ENT clinical trials in PCD. For otologic exacerbations, opinions were similar, however no consensus was reached on the importance of this problem for adults with PCD, primarily due to smaller frequency of acute ear exacerbations in adulthood. The panel also agreed that sinonasal, otologic, and pulmonary exacerbations may occur separately from each other, highlighting again the importance of separate definitions. Responses to key questions about the components of the two definitions are presented in Supplementary Table S2. The combination of new symptoms or worsening of baseline symptoms and of new clinical signs or changes in clinical examination was voted as the best combination of components to define both sinonasal (93%) and otologic (97%) exacerbations. No consensus was reached about including the following components: 1) changes in imaging for sinonasal exacerbations, 2) decision of ENT specialist to treat (for both definitions), and 3) complete resolution of any changes and return to baseline (for both definitions).

Survey 2 included questions on specific elements, particularly symptoms (Supplementary Table S3) and signs (Supplementary Table S4) for the sinonasal and the otologic exacerbation definitions. Agreement was reached for three symptoms and two signs for each definition in this round. Items that achieved 60-79% agreement in survey 2 were discussed again in survey 3. Tables 2 and 3 follow the process of reaching a consensus for the two definitions step by step from survey 2 to survey 4 and the levels of agreement until consensus was reached, or not. Survey 2 also clarified that sinus imaging should not be an absolute requirement for the definition of a sinonasal exacerbation, with main reasoning that it should be restricted for baseline assessment and for complications, and that it entails too much radiation and offers little in case of acute exacerbations (85% agreement).

**Table 2:** Process of reaching consensus for the items included in the definition of a sinonasal exacerbation.

|  | % of agreement |  |  | Included in the definition<br>(% of agreement) |
| --- | --- | --- | --- | --- |
|  | Survey 2 | Survey 3 | Survey 4 |  |
| Patient-reported acute increase in nasal discharge or change in discharge colour | 100 | - | - | major criterion (100) |
| Patient-reported acute pain or sensitivity in the sinus region (i.e. around the nose, eyes, on the cheeks or forehead) | 85 | - | - | major criterion (83) |
| Patient-reported acute blocked nose or worsening in chronic feeling of blocked nose | 92 | - | - | minor criterion (78) |
| Patient-reported acute decreased sense of smell | 69 | 58 | 74 | minor criterion (74) |
| Reduced quality of life evaluated by any sinonasal specific quality of life questionnaire | 73 | 50 | 61 | not included |
| Mucopurulent nasal discharge at examination | 100 | - | - | major criterion (87) |
| Increased mucus production or postnasal drip at examination | 92 | - | - | minor criterion (70) |
| Signs of acute complication (e.g. orbital infection or abscess, meningitis, cerebral infection, cranial nerve palsy) at examination | 72 | 52 | 83 | minor criterion (83) |
| Acute frontonasal or maxillary tenderness at examination | - | - | 65 | not included |
| Doctor's decision to treat, not necessarily with antibiotics but also with increased upper airway clearance or other medication |  | 81 | - | minor criterion (91) |
| Important improvement in symptoms reported by the patient or parent or in clinical findings in case further examination is possible, after a period of at least 14 days | - | 80 | - | minor criterion (74) |

Items that reached $\geq 80\%$ were automatically included in the definition. Items that achieved 60-79% agreement in Survey 2 were discussed again in Survey 3. Items that achieved 50-79% agreement in Survey 3 and newly suggested items by several members were discussed in Survey 4.
At Survey 4, members voted whether items should be considered as major or minor criterion or be included at all. We considered reaching consensus at $\geq 80\%$ agreement for major criteria and $\geq 74\%$ for minor criteria; items with $< 74\%$ agreement were not included at all.

**Table 3:** Process of reaching consensus for the items included in the definition of an otologic exacerbation.

|  | % of agreement |  |  | Included in the definition<br>(% of agreement) |
| --- | --- | --- | --- | --- |
|  | Survey 2 | Survey 3 | Survey 4 |  |
| Patient-reported acute ear sensitivity or pain | 92 | - | - | major criterion (91) |
| Patient-reported acute ear discharge | 92 | - | - | major criterion (91) |
| Patient-reported acute hearing problems or worsening in preexisting hearing problems | 85 | - | - | minor criterion (74) |
| Reported feeling of fullness in the ears | 77 | 58 | 57 | not included |
| Ear discharge at examination | 92 | - |  | major criterion (83) |
| Signs of otitis media in otoscopy (i.e. erythema, collection) | 92 | - |  | major criterion (87) |
| Signs of acute complication (mastoiditis, meningitis, cerebral abscess, facial or other cranial nerve palsy) at examination | 69 | 46 | 78 | minor criterion (78) |
| Impaired hearing tested by pure-tone audiometry | 69 | 62 | 70 | not included |
| Perforated eardrum at examination | 62 | 54 | 43 | not included |
| Horizontal nystagmus at examination | 35 | - | 14 | not included |
| Doctor's decision to treat, not necessarily with antibiotics but also with other medication | - | 88 | 78 | minor criterion (78) |
| Important improvement in symptoms reported by the patient or parent or in clinical findings in case further examination is possible, after a period of 14 days | - | 72 | 70 | not included |

Items that reached $\geq 80\%$ were automatically included in the definition. Items that achieved 60-79% agreement in Survey 2 were discussed again in Survey 3.
Items that achieved 50-79% agreement in Survey 3 and newly suggested items by several members were discussed in Survey 4.
At Survey 4, members voted whether items should be considered as major or minor criterion or be included at all. We considered reaching consensus at $\geq 80\%$ agreement for major criteria and $\geq 74\%$ for minor criteria; items with $< 74\%$ agreement were not included at all.

Survey 3 discussed elements from previous surveys, which had scored highly but not yet reached a consensus on inclusion (Supplementary Table S5). The panel unanimously agreed in this survey to introduce major and minor criteria for both definitions. We reached consent (85%) that all clinical signs or changes seen in clinical examinations included in both definitions should be assessed in relation to previous examinations. In survey 4, participants voted if criteria for which consensus was already reached should be included as major or minor (Tables 2 and 3). Criteria that reached more than 50% but less than 80% agreement in survey 3, were now voted upon including whether to assign as minor or exclude.

Based on discussions that clinical practice may differ substantially from research practices, particularly in the non-PCD ENT specialist, although we originally considered that the definitions would cover also clinical practice, the panel decided (100% agreement) to include the following clarification: “These definitions are aimed to be used in research settings, especially in clinical trials, to define a sinonasal or otologic exacerbation in patients with PCD”. The panel also agreed that a) 3 major or b) 2 major and at least 2 minor criteria are needed to define a sinonasal or otologic exacerbation (Table 4). For sinonasal exacerbation, we reached consensus on three major (reported acute increase in nasal discharge or change in discharge colour, reported acute pain or sensitivity in the sinus region, and mucopurulent nasal discharge at examination) and six minor criteria (reported acute blocked nose or worsening in chronic feeling of blocked nose, reported acute decreased sense of smell, increased mucus production or postnasal drip at examination, signs of acute complication at examination, doctoŕs decision to treat, and important improvement in symptoms or clinical findings after a period of at least 14 days). For an otologic exacerbation, we reached consensus on 4 major (reported acute ear sensitivity or pain, reported acute ear discharge, ear discharge at examination, and sign of otitis media in otoscopy) and 3 minor (reported acute hearing problems/worsening in preexisting hearing problems, signs of acute complications at examination, and doctor’s decision to treat). Major criteria were decided on at least 80% consensus and minor on at least 74%, which was agreed in the panel (Tables 2 and 3). Lastly, the panel highlighted that no criterion was an absolute requirement for either definition (Table 4).

**Table 4:**
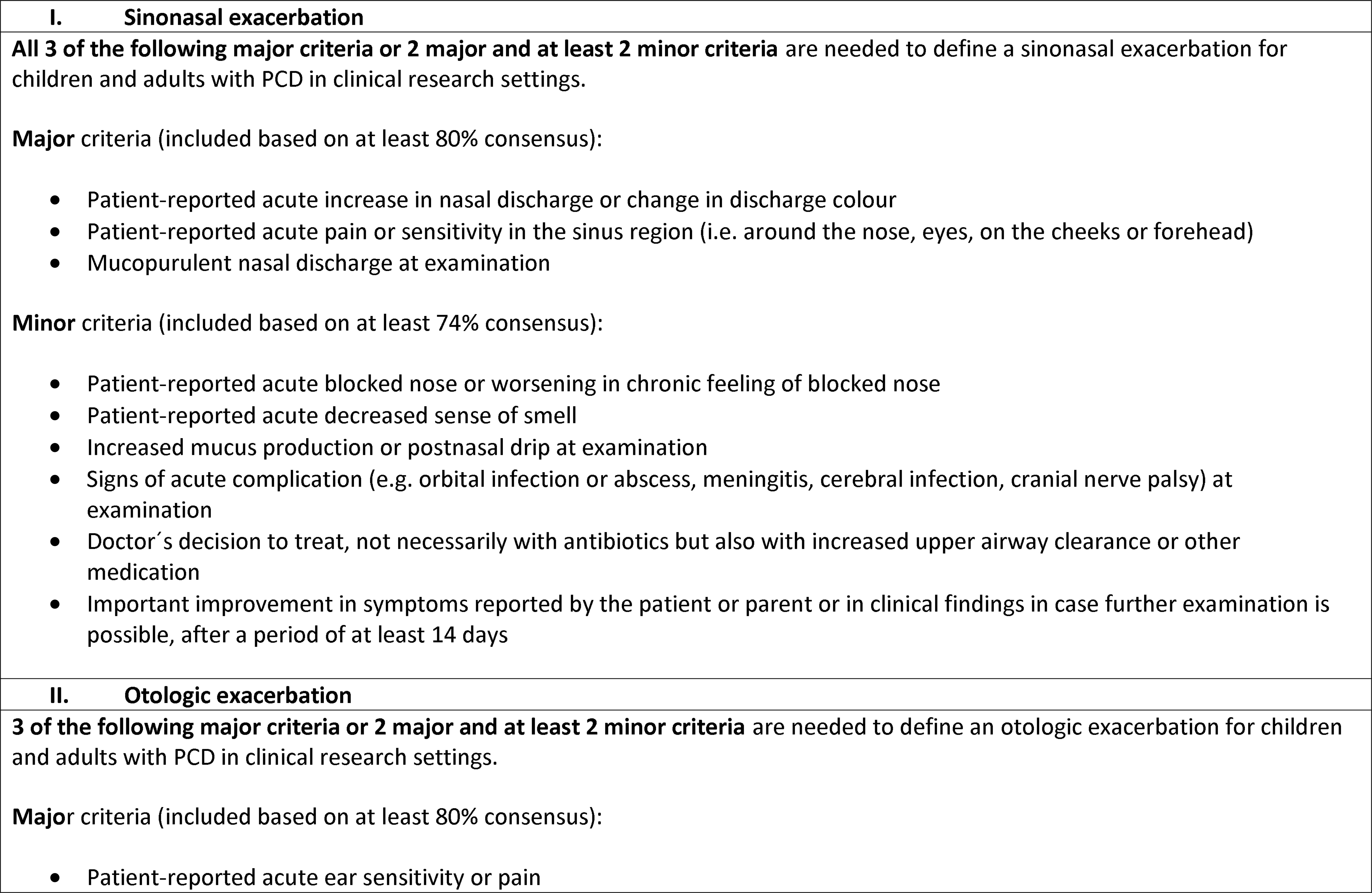
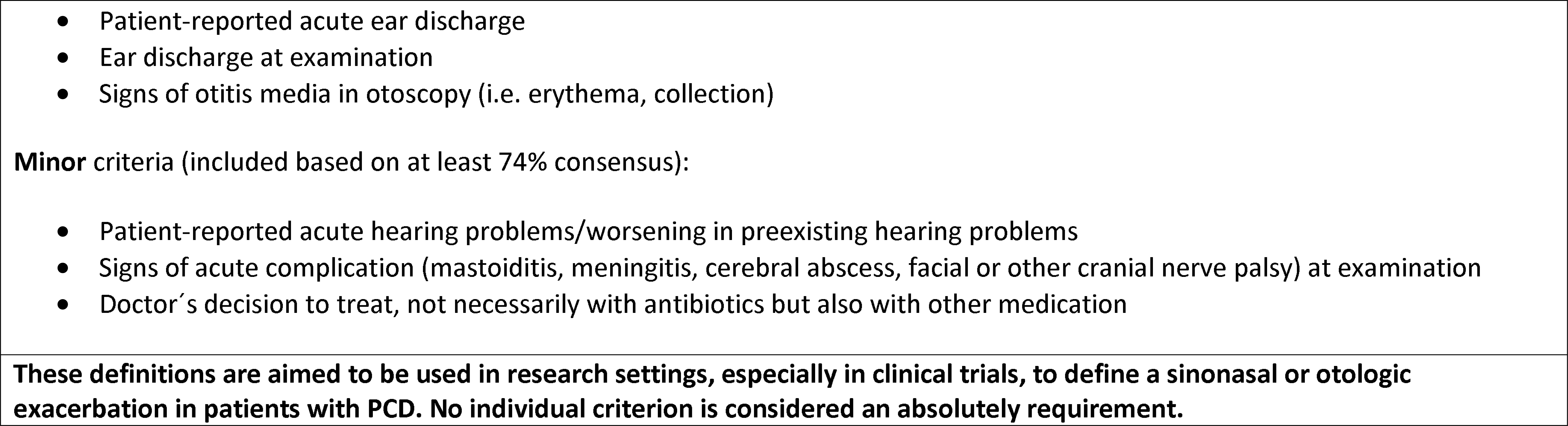
Definitions of a sinonasal and an otologic exacerbation for children and adults with primary ciliary dyskinesia (PCD) participating in clinical research.

## Discussion

An international panel of ENT specialists, pulmonologists, healthcare professionals, and people with PCD, agreed on consensus definitions of sinonasal and otologic exacerbations in children and adults with PCD to be used in research, especially in clinical trials. This effort followed similar approach to the one used to develop a consensus definition of pulmonary exacerbations in PCD.[18] Although upper and lower airway disease in PCD should be managed in unison and exacerbations often occur simultaneously, or expand to involve the whole airways, our panel agreed that exacerbations from the nose, the sinuses, and the ears require separate definitions.[54] They can occur individually and have different characteristics. Both are an important problem in children with PCD and whilst in adults sinonasal exacerbations remain a major issue, otologic exacerbations are less common.

Main strengths of the study were the international and multidisciplinary nature of the panel, and the inclusion of patients and parents of children with PCD, together with the added group of patient volunteers. We performed a thorough systematic review of the literature, expanding our search also to other conditions, such as other types of CRS, which have similarities with PCD. We retained a high panel response rate throughout the study. Although the panel considered originally developing definitions that would also be used for clinical practice, we agreed during the process that this would not be feasible. However simple, clinical outcomes need to be very clearly defined, while for clinical practice a decision for an exacerbation might be needed to be taken only based on reported symptoms, often without any examination.

Our panel discussed thoroughly whether existing definitions specifically for exacerbation of CRS, could be used also for children and adults with PCD, without the need to develop disease-specific definitions. We considered all available definitions (Table 1), particularly the latest European Position Paper on Rhinosinusitis and Nasal Polyps (EPOS) that defined acute exacerbation of CRS as worsening of symptom intensity with return to baseline CRS symptom intensity, often after intervention with corticosteroids and or antibiotics.[36] We reached consensus that none of them fully covered the purpose of a PCD-specific definition, although they highlighted important components which we then discussed. In particular, the panel members agreed on the need to include in the definition PCD-specific signs seen at simple examination. We found no eligible definitions that could be used as a starting point for otologic exacerbations.

Throughout the process, our panel highlighted the need to select elements which could be assessed easily, in different settings and would not require complex ENT examination or a specialist with expertise in PCD to assess them. Most criteria refer to symptoms, or signs that can be observed in simple clinical examination, the most complex assessment included being otoscopy. Panel members agreed that patients with PCD often underestimate their upper airway symptoms, which are non-specific and to which they grown accustomed over time, highlighting the need to also consider simple signs in the definitions.[4, 7, 55, 56] This was also shown in a recent study from the ENT Prospective International Cohort of PCD Patients (EPIC-PCD) that reported a lack of correlation between sinonasal and otologic symptoms with objective measurements.[57, 58]

Two components that required long discussions and voting rounds were doctor’s decision to treat and the need for improvement of the symptoms and signs. In both definitions, decision to treat was included as minor criterion since it could occur regardless of an exacerbation (e.g. detection of Pseudomonas aeruginosa in a routine nasal or ear sample). The panel clarified that treatment should not only refer to need for antibiotics but also other medication or management practices such as upper airway clearance. Return to baseline was a term that was not found agreeable to most panel members. Even though improvement in symptoms and signs, where follow-up examination is possible, was included as a minor criterion for the sinonasal definition, participants agreed that it is difficult to measure improvement as deterioration is partly expected due to the chronic nature of the disease. In case of acute ear exacerbations especially, this was not considered possible, and it was not included at all in the definition.

This initiative was developed in the framework of the BEAT-PCD ERS clinical research collaboration (https://beat-pcd.squarespace.com), as part of our efforts to define and promote the use of reliable clinical outcome measures for PCD trial and clinical research.[20] Evidence-base for PCD treatment is small, and there are no trials which have accessed specifically management of the upper airways. Identifying the most suitable clinical and patient-reported outcomes to be used as endpoints focused on the upper and lower airways, was one of the top priorities related to PCD research identified recently by experts in the field. As more trials are needed to improve care of PCD and new potential therapies are in the pipeline, these definitions might offer a useful outcome measure in different research settings. It is important to use and validate them in future studies and trials, to assess their usability together with other potential outcomes.

## Supporting information

eDelphi survey Supplementary material 1-4

## Acknowledgments

We would like to thank Jeanette Boyd from the European Lung Foundation (ELF) and the group of people affected by PCD who have supported this project by participating in the eDelphi surveys and providing feedback outside of the consensus expert panel, namely: Tanja Hedberg (Sweden), Tamar Makhatadze (Georgia), Nina Peters (UK), Poonam Sodha (UK) and Peter van Baalen (Netherlands).

## Author Contributions

M Goutaki and JF Papon developed the concept and designed the study. YT Lam performed the literature search. M Goutaki managed the study, designed the eDelphi surveys, analysed the data, and drafted the manuscript. All authors contributed to the surveys, interpreted results, and revised critically the manuscript. M Goutaki and JF Papon take final responsibility for all content.

## Data availability

All data generated for this project was made available in the manuscript display items or supplementary information.

