## Supplementary material for "Definition of sinonasal and otologic exacerbation in patients with primary ciliary dyskinesia - an expert consensus": eDelphi survey Supplementary material 1-4

**Definition of ENT exacerbations in PCD Consensus eDelphi Survey 1**

**Please read before completing the survey:**

The aim of this project is to reach a consensus for defining exacerbations from the upper airways in people with PCD at the end of this eDelphi survey process.

This definition is meant to be used for clinical trials and other research and for clinical practice. At the first panel meeting, we concluded this standardised definition was missing and we decided unanimously to produce two separate definitions: one for sinonasal exacerbations (from the nose and sinuses) and one for otologic exacerbations (from the ears).

At the second panel meeting, we presented: 1) a summary of first opinions from the ENT experts about these definitions (anonymously) and 2) the results of the literature review we performed separately for nose/ sinuses (including the EPOS definition for non-PCD rhinosinusitis which was highlighted by several members) and ears. Based on both experts’ first opinions and existing literature, we have identified the following 4 main points as important components for both definitions:

1) new symptoms/ worsening of baseline symptoms or quality of life
2) clinical signs/changes in clinical examinations
3) physician´s decision to treat/change management
4) complete resolution of changes/return to baseline

In this first eDelphi survey we include questions about these main components so we can reach a consensus which should be included in each definition. As the eDelphi surveys go on, we will go into more detail about the chosen components to decide e.g. which symptoms or which signs (if symptoms or signs are selected respectively).

Your replies will remain anonymous to the panel and only the moderating team will have access to your identifying information.

Please make sure you reply all questions.

I. **Few questions about your background and your relationship to PCD**

- Your name

(text box)

- Country you work/live in

(text box)

- Are you (please select one of the following categories):

ENT specialist

Pulmonologist (paediatric or adult)

Nurse

Physiotherapist

Epidemiologist

Person with PCD or parent of child with PCD

- How many years do you have experience with PCD?
- If you are an ENT specialist do you manage?

children with PCD

adults with PCD

both children and adults

not an ENT specialist

- If you are ENT specialist, do you perform?

ear surgery

sinonasal surgery

II. **Sinonasal exacerbation in people with PCD (linked to the nose and sinuses)**

- What is your opinion about each of the following statements?

(choose of the following: strongly disagree, disagree, neither disagree nor agree, agree, strongly agree)

Exacerbations from the nose and sinuses are an important problem for adults with PCD.

Exacerbations from the nose and sinuses are an important problem for children with PCD.

Exacerbations from the nose and sinuses impact the quality of life of people with PCD.

Exacerbations from the nose and sinuses always occur together with exacerbations from the ears in people with PCD.

Exacerbations from the nose and sinuses always occur together with exacerbations from the lungs in people with PCD.

Exacerbations from the nose and sinuses are not always dependent on upper airway disease baseline severity in people with PCD.

Exacerbations from the nose and sinuses can be an important outcome measure for ENT clinical trials in PCD.

- Do you have any other relevant comments on the importance of sinonasal exacerbations?

(text box)

- In your opinion which of the following components can provide the best definition for an exacerbation from the nose and sinuses:

(choose of the following: strongly disagree, disagree, neither disagree nor agree, agree, strongly agree)

New symptoms/ worsening of baseline symptoms from the nose and sinuses, in isolation

Clinical signs/changes in clinical examinations of the nose and sinuses (e.g. nasal endoscopy), in isolation

Changes in imaging (CT or other) of the nose and sinuses in isolation

Decision of ENT specialist to treat, in isolation

Combination of symptoms and changes in clinical examination

Combination of symptoms and decision of ENT specialist to treat

Combination of symptoms and changes in imaging

Combination of changes in clinical examinations and decision of ENT specialist to treat

Combination of changes in clinical examinations and changes in imaging

Changes in imaging and decision of ENT specialist to treat

Combination of symptoms and changes in clinical examinations and decision of ENT specialist to treat

Combination of symptoms and changes in clinical examinations and imaging

Combination of symptoms and changes in imaging and decision of ENT specialist to treat

Combination of symptoms, changes in clinical examinations and imaging and decision of ENT specialist to treat

- What is your opinion about the following statement:

(choose of the following: strongly disagree, disagree, neither disagree nor agree, agree, strongly agree)

Complete resolution of any changes/return to baseline is an important element for the definition of exacerbations from the nose and sinuses in PCD

- Do you have anything to add regarding the general components for the definition of sinonasal exacerbations (remember, we will go into detail in the next surveys) or do you have any comment explaining your replies?

(text box)

III. **Otologic exacerbation in people with PCD (linked to the ears)**

- What is your opinion about each of the following statements?

(choose of the following: strongly disagree, disagree, neither disagree nor agree, agree, strongly agree)

Exacerbations from the ears are an important problem for adults with PCD.

Exacerbations from the ears are an important problem for children with PCD.

Exacerbations from the ears impact the quality of life of people with PCD.

Exacerbations from the ears always occur together with exacerbations from the nose and sinuses in people with PCD.

Exacerbations from the ears always occur together with exacerbations from the lungs in people with PCD.

Exacerbations from the ears are not always dependent on upper airway disease baseline severity in people with PCD.

Exacerbations from the ears can be an important outcome measure for ENT clinical trials in PCD.

- Do you have any other relevant comments on the importance of otologic exacerbations?

(text box)

- In your opinion which of the following components can provide the best definition for an exacerbation from the ears:

(choose of the following: strongly disagree, disagree, neither disagree nor agree, agree, strongly agree)

New symptoms/ worsening of baseline symptoms from the ears, in isolation

Clinical signs/changes in clinical examinations of the ears (e.g. otoscopy, audiometry), in isolation

Decision of ENT specialist to treat, in isolation

Combination of symptoms and changes in clinical examinations

Combination of symptoms and decision of ENT specialist to treat

Combination of changes in clinical examinations and decision of ENT specialist to treat

Combination of symptoms and changes in clinical examinations and decision of ENT specialist to treat

- What is your opinion about the following statement:

(choose of the following: strongly disagree, disagree, neither disagree nor agree, agree, strongly agree)

Complete resolution of any changes/return to baseline is an important element for the definition of exacerbations from the ears in PCD.

- Do you have anything to add regarding the general components for the definition of otologic exacerbations (remember, we will go into detail in the next surveys) or do you have any comment explaining your replies?

(text box)

**Thank you for completing this survey, please contact** **if you have any questions.**

**Definition of ENT exacerbations in PCD Consensus eDelphi Survey 2**

**Please read before completing the survey:**

We would like to remind you that the aim of this project is to reach a consensus for defining exacerbations from the upper airways in people with PCD at the end of this eDelphi survey process.

This definition is meant to be used for clinical trials and other research and for clinical practice. We decided unanimously to produce two separate definitions: one for sinonasal exacerbations and one for otologic exacerbations.

With the first survey, we assessed opinions about the importance of sinonasal and otologic exacerbations for people with PCD and about the components that should be included in the exacerbation definitions. We achieved 100% participation of panel and patient advisory group members in the first survey.

After evaluating all responses and comments, we have now developed this second survey that includes questions to help us get closer to a consensus for the definition of sinonasal and otologic exacerbations. This second survey is longer as it is going into more details about the components highlighted in the first survey and will help decide on all included elements. A next survey based on these results will help to clarify open questions.

Your replies will remain anonymous to the panel and only the moderating team will have access to your identifying information.

IV. **Few questions about your background and your relationship to PCD**

- Name

(text box)

- Country you work/live in

(text box)

V. **Sinonasal exacerbation in people with PCD (linked to the nose and sinuses)**

**In the first survey, we reached consensus (more than 80% agreed or strongly agreed) on the following statements about the importance of sinonasal exacerbations:**

1. Exacerbations from the nose and sinuses are an important problem for adults with PCD.
2. Exacerbations from the nose and sinuses are an important problem for children with PCD.
3. Exacerbations from the nose and sinuses impact the quality of life of people with PCD.
4. Exacerbations from the nose and sinuses can be an important outcome measure for ENT clinical trials in PCD.

**We also reached consensus about sinonasal exacerbations occurring also separately from pulmonary or otologic exacerbations.**

- Do you have any additional comments about the importance of sinonasal exacerbations? (text box)

**In the first survey, the respondents said that the combination of new symptoms/ worsening of baseline symptoms and of clinical signs/ changes in clinical examination is the best combination of components to define sinonasal exacerbations.**

**Change in symptoms**

- Should changes in the following symptoms be included as an indication of a sinonasal exacerbation?

(5 level system: strongly disagree, disagree, neither disagree nor agree, agree, strongly agree)

Fever/Temperature above 38°C

Blocked nose

Increase in nasal discharge or change in discharge colour

Decreased sense of smell

Pain/sensitivity in the sinus region (i.e. around the nose, eyes, on the cheeks or forehead)

Headache

Malaise, fatigue, tiredness

Increased cough

Sore throat

Bad breath

Bad sleep

Reduced quality of life (QoL) evaluated by any sinonasal specific QoL questionnaire

- Are there any other symptoms, which should be included as an indication of a sinonasal exacerbation? Please explain (text box)
- Please rank the 3 most important symptoms (from the list above or any additional) for inclusion in the definition

**Changes in clinical examinations**

- Should the following clinical signs/changes in examinations be included as an indication of a sinonasal exacerbation? (Note: this would mean that the agreed upon examinations should be performed in all people with PCD to diagnose a sinonasal exacerbation) (yes or no)

Erythema or oedema of nasal mucosa

Mucopurulent nasal discharge

Increased mucus production or postnasal drip

Swelling of inferior turbinates

Appearance or swelling of nasal polyps

Positive microbiology culture from nasal or sinus discharge

Reduced sense of smell measured by any olfactory test (smell test)

Signs of acute complication (e.g. orbital infection/abscess, meningitis, cerebral infection, cranial nerve palsy)

Negative allergy tests (to exclude allergic causes)

Blood tests for indications of infection (e.g. neutrophils)

- Are there any other findings of clinical examinations, which should be included as an indication of a sinonasal exacerbation? Please explain (text box)
- Please rank the 3 most important examination findings (from the list above or any additional) for inclusion in the definition

**In the first round of the survey, there was no consensus that changes in imaging should be a requirement for the definition of a sinonasal exacerbation. However, a majority indicated that it might contribute in combination with changes in symptoms and clinical examinations.**

- Should a change in sinus imaging be an absolute requirement for the definition of a sinonasal exacerbation and therefore should it be performed in all people with PCD to diagnose a sinonasal exacerbation?

(2 choices: Agree, Disagree)

- Please explain your vote above (text box)

**In the first round of the survey, there was no consensus that decision of ENT specialist to treat** **should be a requirement for the definition of a sinonasal exacerbation. However, a majority indicated that it might contribute in combination with changes in symptoms and clinical examinations.**

- Should the decision of ENT specialist to treat be an absolute requirement for the definition of a sinonasal exacerbation?

(2 choices: Agree, Disagree)

- Please explain your vote above (text box)
- If the decision of an ENT specialist to treat is considered for the definition, which of the following treatments should be considered (one choice only possible):

Prescription of antibiotics

Changes in non-antibiotic management (e.g. upper airways clearance or other medication)

Any of the two

None, it should not be part of the definition

**In the first round of the survey, there was no consensus that complete resolution of any changes/return to baseline should be a component for the definition of sinonasal exacerbation. A majority indicated that it might be important to include. Others stated that complete resolution is not possible in many cases so it cannot be an absolute requirement for the definition.**

- Should complete resolution and return to baseline be an absolute requirement for the definition of a sinonasal exacerbation?

(2 choices: Agree, Disagree)

- Should an important improvement in symptoms/clinical findings after a period of time be an absolute requirement for the definition of a sinonasal exacerbation?

(2 choices: Agree, Disagree)

- Please explain your votes above (text box)
- Do you have any further comments about the definition of sinonasal exacerbations? (text box)

VI. **Otologic exacerbation in people with PCD (linked to the ears)**

**In the first survey, we reached consensus (more than 80% agreed or strongly agreed) on the following statements about the importance of otologic exacerbations:**

1. Exacerbations from the ears are an important problem for children with PCD.
2. Exacerbations from the ears impact the quality of life of people with PCD.
3. Exacerbations from the ears can be an important outcome measure for ENT clinical trials in PCD.

Respondents did not reach a consensus that otologic exacerbations are an important problem for adults with PCD but a majority voted that it is.

- In your opinion, are exacerbations from the ears an important problem for adults with PCD?

(2 choices: Agree, Disagree)

- Please explain your vote above, especially if you disagree with the statement (text box)

**We also reached consensus about otologic exacerbations occurring also separately from pulmonary or sinonasal exacerbations.**

- Do you have any additional comments about the importance of otologic exacerbations?

(text box)

**In the first survey, the respondents said that the combination of new symptoms/ worsening of baseline symptoms and of clinical signs/ changes in clinical examination is the best combination of components to define otologic exacerbations. Decision of an ENT specialist to treat in addition to this combination, was the second most popular choice (also reached consensus).**

**Change in symptoms**

- Should changes in the following symptoms be included as an indication of an otologic exacerbation?

(5 level system: strongly disagree, disagree, neither disagree nor agree, agree, strongly agree)

Fever/Temperature above 38°C

Ear/sensitivity pain

Ear discharge

Tinnitus or ringing in the ears

A feeling of fullness in the ears

Hearing problems

Difficulty for communication

Malaise, fatigue, tiredness

Headache

Dizziness, vertigo

- Are there any other symptoms, which should be included as an indication of an otologic exacerbation? Please explain (text box)
- Please rank the 3 most important symptoms (from the list above or any additional) for inclusion in the definition

**Changes in clinical examinations**

- Should the following clinical signs/changes in examinations be included as an indication of an otologic exacerbation? (yes or no)

Perforated eardrum

Ear discharge

Nystagmus

Signs of otitis media in otoscopy (i.e. erythema, collection)

Pathologic tympanometry findings

Signs of acute complication (mastoiditis, meningitis, cerebral abscess, facial or other cranial nerve palsy…)

Impaired hearing tested by pure-tone audiometry (air and bone-conduction)

Positive microbiology culture from ear discharge

Blood tests for indications of infection (e.g. neutrophils)

- Are there any other findings of clinical examinations, which should be included as an indication of an otologic exacerbation? Please explain (text box)
- Please rank the 3 most important examination findings (from the list above or any additional) for inclusion in the definition

**Decision of ENT specialist to treat**

- Should the decision of ENT specialist to treat be an absolute requirement for the definition of an otologic exacerbation?

(5 level system: strongly disagree, disagree, neither disagree nor agree, agree, strongly agree)

- If the decision of an ENT specialist to treat is considered for the definition, which of the following treatments should be considered (one choice only possible):

Prescription of antibiotics (either local or systemic)

Changes in non-antibiotic management (e.g. ear antiseptic or other medication)

Any of the two

None, it should not be part of the definition

**In the first round of the survey, there was no consensus that complete resolution of any changes/return to baseline should be a component for the definition of otologic exacerbation. A majority indicated that it might be important to include. Others stated that complete resolution is not possible in many cases so it cannot be an absolute requirement for the definition.**

- Should complete resolution and return to baseline be an absolute requirement for the definition of an otologic exacerbation?

(2 choices: Agree, Disagree)

- Should an important improvement in symptoms/clinical findings after a period of time be an absolute requirement for the definition of an otologic exacerbation?

(2 choices: Agree, Disagree)

- Please explain your votes above (text box)
- Do you have any further comments about the definition of otologic exacerbations? (text box)

**Thank you for completing this survey, please contact** **if you have any questions.**

**Definition of ENT exacerbations in PCD Consensus eDelphi Survey 3**

**Please read before completing the survey:**

We would like to remind you that the aim of this project is to reach a consensus for defining exacerbations from the upper airways in people with PCD at the end of this eDelphi survey process.

This definition is meant to be used for clinical trials and other research and for clinical practice.

With the first survey, we assessed opinions about the importance of sinonasal and otologic exacerbations for people with PCD and about the components that should be included in the exacerbation definitions and with the second we went into the specific elements for the relevant components where we reached consensus. We achieved 100% participation of the panel and a very good participation from the patient advisory group members in both surveys.

After evaluating all responses and comments, we have now developed this third survey that clarifies opinions on the elements that were voted on in the previous one and aims to finalise all components and elements for the two definitions. This third survey is also long but allow us to decide on all the elements included in the two definitions.

A next last survey based on these results will help to clarify all remaining open questions. We aim to organise a group meeting after analysing the 3^rd^ survey results and before drafting the 4^th^ survey to make sure we discuss all important points that needs to be included in survey 4.

Your replies will remain anonymous to the panel and only the moderating team will have access to your identifying information.

VII. **Few questions about your background and your relationship to PCD**

- Name

(text box)

- Country you work/live in

(text box)

VIII. **Sinonasal exacerbation in people with PCD (linked to the nose and sinuses)**

**Change in symptoms**

**In the second survey, we reached consensus (more than 80% agreed or strongly agreed) that the following 3 symptoms (or worsening in them) should be included as an indication of a sinonasal exacerbation. These 3 symptoms were also ranked as the 3 most important by the majority of participants with great difference in votes from all others.**

- Increase in nasal discharge or change in discharge colour
- Pain/sensitivity in the sinus region (i.e. around the nose, eyes, on the cheeks or forehead)
- Blocked nose

1. What is your opinion about the inclusion of changes in clinical symptoms in the definition (choose one):

- A patient should have ALL symptoms that we will include in the definition to be considered as having a sinonasal exacerbation.
- We should include a list of relevant important symptoms of which a patient should have a combination but NOT ALL, to be considered as having a sinonasal exacerbation.

**In the second round of the survey, there was no consensus about the inclusion of the following symptoms in the definition as an indication of a sinonasal exacerbation. However, they were voted as important by 55-75% of participants and were ranked among the 3 most important symptoms by few.**

2. Should reduced quality of life evaluated by any sinonasal specific quality of life (QoL) questionnaire be included in the definition as an indication of a sinonasal exacerbation?

(2 choices: Agree, Disagree)

3. Please explain your vote above (text box)
4. Should decreased sense of smell be included in the definition as an indication of a sinonasal exacerbation?

(2 choices: Agree, Disagree)

5. Please explain your vote above (text box)
6. Should headaches be included in the definition as an indication of a sinonasal exacerbation?

(2 choices: Agree, Disagree)

7. Please explain your vote above (text box)
8. Should bad sleep be included in the definition as an indication of a sinonasal exacerbation?

(2 choices: Agree, Disagree)

9. Please explain your vote above (text box)
10. Should cough be included in the definition as an indication of a sinonasal exacerbation?

(3 choices: Agree for all, Agree only for children, Disagree)

11. Please explain your vote above (text box)
12. Are there any other symptoms not mentioned so far in this survey, which you find very important for a sinonasal exacerbation and you would like to discuss/put up for voting again?

(Text box)

**Changes in clinical examinations**

**In the second survey, we reached consensus (more than 80% agreed or strongly agreed) that the following 2 clinical signs/changes in examinations be included as an indication of a sinonasal exacerbation. These 2 clinical signs/changes in examinations were also ranked as the 2 most important by the majority of participants with great difference in votes from all others.**

- Mucopurulent nasal discharge
- Increased mucus production or postnasal drip

13. What is your opinion about the inclusion of clinical signs/changes in examinations in the definition (choose one):

- A patient should have ALL clinical signs/changes in examinations that we will include in the definition to be considered as having a sinonasal exacerbation.
- We should include a list of relevant important clinical signs/changes in examinations of which a patient should have a combination but NOT ALL, to be considered as having a sinonasal exacerbation.

**In the second round of the survey, there was no consensus about the inclusion of the following clinical signs/changes in examinations in the definition as an indication of a sinonasal exacerbation. However, they were voted as important by 55-75% of participants and were ranked among the 3 most important clinical signs/changes in examinations by few.**

14. Should signs of acute complication (e.g. orbital infection/abscess, meningitis, cerebral infection, cranial nerve palsy) be included in the definition as an indication of a sinonasal exacerbation?

(2 choices: Agree, Disagree)

15. Please explain your vote above (text box)
16. Should appearance or swelling of nasal polyps be included in the definition as an indication of a sinonasal exacerbation?

(2 choices: Agree, Disagree)

17. Please explain your vote above (text box)
18. Should erythema or oedema of nasal mucosa be included in the definition as an indication of a sinonasal exacerbation?

(2 choices: Agree, Disagree)

19. Please explain your vote above (text box)
20. Are there any other clinical signs/changes in examinations not mentioned so far in this survey, which you find very important for a sinonasal exacerbation and you would like to discuss/put up for voting again?

(Text box)

**In the second round of the survey, we reached consensus (more than 80% agreed or strongly agreed) that changes in imaging should not be a requirement for the definition of a sinonasal exacerbation.**

**Decision to treat and resolution of changes**

**In the second round of the survey, there was no consensus about including or not including decision of ENT specialist to treat** **as a requirement for the definition of a sinonasal exacerbation. However, a majority indicated that it might contribute in combination with changes in symptoms and clinical examinations.**

21. What is your opinion on the following statements related to sinonasal exacerbations

(2 choices for all: Agree, Disagree)

- Decision to treat might be subjective and should not be included especially for the use of the definition in clinical trials.
- Decision to treat is based on the changes of symptoms and clinical examinations and does not need to be included in the definition in addition to those.
- Decision to treat is more relevant for the use of the definition in clinical practice, however we should include other physicians e.g. paediatrician, general practitioner as in reality, not all patients need to visit an ENT to diagnose a sinonasal exacerbation.
- Decision to treat not necessarily with antibiotics but also with increased upper airways clearance or other medication, could be included in the definition as a less important point (e.g. possibly a minor criterion).

22. Do you have any comments about these statements?

(text box)

**In the second round of the survey, we reached consensus that complete resolution of any changes and return to baseline should not an absolute requirement for the definition of sinonasal exacerbation. However, a majority of participants voted that an important improvement in symptoms/clinical findings after a period of time should be an absolute requirement for the definition of a sinonasal exacerbation.**

23. What is your opinion on the following statements:

- An important improvement in symptoms/clinical findings after a period of time should be included in the definition of a sinonasal exacerbation as a less important point (e.g. possibly a minor criterion).
- The absence of improvement in symptoms/clinical findings does not eliminate the diagnosis of acute sinonasal exacerbation.

(2 choices: Agree, Disagree)

24. Please explain your vote above (text box)
25. Do you have any further comments about the definition of sinonasal exacerbations? (text box)
IX. **Otologic exacerbation in people with PCD (linked to the ears)**

**In the second survey, we did not reach a consensus that otologic exacerbations are an important problem for adults, however many participants commented that they only disagreed because otologic exacerbations are less common in adults.**

26. What is your opinion on the following statement:

- Exacerbations from the ears are less common than chronic problems in adults with PCD, however when they occur, they can be an important problem.

(2 choices: Agree, Disagree)
27. Please explain your vote above (text box)

**Change in symptoms**

**In the second survey, we reached consensus (more than 80% agreed or strongly agreed) that the following 3 symptoms (or worsening in them) should be included as an indication of an otologic exacerbation. These 3 symptoms were also ranked as the 3 most important by the majority of participants with great difference in votes from all others.**

- Ear sensitivity/pain
- Ear discharge
- Hearing problems

28. What is your opinion about the inclusion of changes in clinical symptoms in the definition (choose one):

- A patient should have ALL symptoms that we will include in the definition to be considered as having an otologic exacerbation.
- We should include a list of relevant important symptoms of which a patient should have a combination but NOT ALL, to be considered as having an otologic exacerbation.

**In the second round of the survey, there was no consensus about the inclusion of the following symptom in the definition as an indication of an otologic exacerbation. However, it was voted as important by 77% of participants and was ranked among the 3 most important symptoms by many.**

29. Should a feeling of fullness in the ears be included in the definition as an indication of an otologic exacerbation?

(2 choices: Agree, Disagree)

30. Please explain your vote above (text box)
31. Are there any other symptoms not mentioned so far in this survey, which you find very important for an otologic exacerbation and you would like to discuss/put up for voting again?

(Text box)

**Changes in clinical examinations**

**In the second survey, we reached consensus (more than 80% agreed or strongly agreed) that the following two clinical signs/changes in examinations be included as an indication of an otologic exacerbation. These 2 clinical signs/changes in examinations were also ranked as the 2 most important by the majority of participants with great difference in votes from all others.**

- Ear discharge
- Signs of otitis media in otoscopy (i.e. erythema, collection)

32. What is your opinion about the inclusion of clinical signs/changes in examinations in the definition (choose one?):

- A patient should have ALL clinical signs/changes in examinations that we will include in the definition to be considered as having an otologic exacerbation.
- We should include a list of relevant important clinical signs/changes in examinations of which a patient should have a combination but NOT ALL, to be considered as having an otologic exacerbation.

**In the second round of the survey, there was no consensus about the inclusion of the following clinical signs/changes in examinations in the definition as an indication of an otologic exacerbation. However, they were voted as important by 50-75% of participants and were ranked among the 3most important clinical signs/changes in examinations by few.**

33. Should signs of acute complication (mastoiditis, meningitis, cerebral abscess, facial or other cranial nerve palsy) be included in the definition as an indication of an otologic exacerbation?

(2 choices: Agree, Disagree)

34. Please explain your vote above (text box)
35. Should impaired hearing tested by pure-tone audiometry be included in the definition as an indication of an otologic exacerbation?

(2 choices: Agree, Disagree)

36. Please explain your vote above (text box)
37. Should a perforated eardrum be included in the definition as an indication of an otologic exacerbation?

(2 choices: Agree, Disagree)

38. Please explain your vote above (text box)
39. Should pathologic tympanometry be included in the definition as an indication of an otologic exacerbation?

(2 choices: Agree, Disagree)

40. Please explain your vote above (text box)
41. Should pathologic pneumatic otoscopy be included in the definition as an indication of an otologic exacerbation, particularly when tympanometry is not available?

(2 choices: Agree, Disagree)

42. Please explain your vote above (text box)
43. Are there any other clinical signs/changes in examinations not mentioned so far in this survey, which you find very important for an otologic exacerbation and you would like to discuss/put up for voting again?

(Text box)

**Decision to treat and resolution of changes**

**In the second round of the survey, there was no consensus about including or not including decision of ENT specialist to treat** **as a requirement for the definition of an otologic exacerbation. However, a majority indicated that it might contribute in combination with changes in symptoms and clinical examinations.**

44. What is your opinion on the following statements related to otologic exacerbations?

(2 choices for all: Agree, Disagree)

- Decision to treat might be subjective and should not be included especially for the use of the definition in clinical trials.
- Decision to treat is based on the changes of symptoms and clinical examinations and does not need to be included in the definition in addition to those.
- Decision to treat is more relevant for the use of the definition in clinical practice, however we should include other physicians e.g. paediatrician, general practitioner as in reality, not all patients need to visit an ENT to diagnose an otologic exacerbation.
- Decision to treat not necessarily with antibiotics but also with other medication, could be included in the definition as a less important point (e.g. possibly a minor criterion).

45. Do you have any comments about these statements?

(text box)

**In the second round of the survey, we reached consensus that complete resolution of any changes and return to baseline should not an absolute requirement for the definition of otologic exacerbation. However, a majority of participants voted that an important improvement in symptoms/clinical findings after a period of time should be an absolute requirement for the definition of an otologic exacerbation.**

46. What is your opinion on the following statements:

- An important improvement in symptoms/clinical findings after a period of time should be included in the definition as a less important point (e.g. possibly a minor criterion), as it is not always possible (e.g. in case of a perforated eardrum)
- The absence of improvement in symptoms/clinical findings does not eliminate the diagnosis of acute otologic exacerbation.

(2 choices: Agree, Disagree)

47. Please explain your vote above (text box)
48. Do you have any further comments about the definition of otologic exacerbations? (text box)
X. **These last questions relates to BOTH definitions (sinonasal and otologic):**
49. Do you agree with the idea of introducing major and minor criteria to the definition (to decide which belong to major or minor category in the last survey)?

(2 choices: Agree, Disagree)

**In the second round of the survey, several participants highlighted the need to assess all clinical examinations findings in comparison to previous (baseline) evaluations.**

50. Should we include the following statement as comment to our definitions related to clinical signs and changes in examinations?

All clinical signs/changes to clinical examinations should be assessed in relation to previous examinations.

(2 choices: Agree, Disagree)

51. Please explain your vote above (text box)

**Thank you for completing this survey and for participating to this project, please contact** **if you have any questions.**

**Definition of ENT exacerbations in PCD Consensus eDelphi Survey 4**

**Please read carefully before completing the survey:**

This is the 4^th^ and final survey.

We would like to remind you that the aim of this project is to reach a consensus for defining exacerbations of the upper airways in people with PCD at the end of this eDelphi survey process.

With the previous surveys, we assessed opinions about the importance of sinonasal and otologic exacerbations for people with PCD. We selected the components that should be included in the exacerbation definitions as well as the specific elements for the relevant components where we reached consensus.

**In the third survey, 100% of participants agreed to introduce major and minor criteria to the definitions. In this fourth and last survey, we will decide which specific elements will be considered as major and which as minor for each definition and will clarify any remaining important points.**

Your replies will remain anonymous to the panel and only the moderating team will have access to your identifying information.

XI. **Few questions about your background and your relationship to PCD**

- Name

(text box)

- Country you work/live in

(text box)

XII. **Sinonasal exacerbation in people with PCD (linked to the nose and sinuses)**

**Change in symptoms:** In the previous surveys, we reached consensus (96% agreed) that **the definition should include a list of relevant important symptoms of which a patient should have a combination** but NOT ALL, to be considered as having a sinonasal exacerbation. We also reached consensus (more than 80% agreed or strongly agreed) that the following 3 symptoms (or worsening in them) should be included as an indication of a sinonasal exacerbation: **i) increase in nasal discharge or change in discharge colour, ii) pain/sensitivity in the sinus region (i.e. around the nose, eyes, on the cheeks or forehead) or iii) blocked nose.**

**Changes in clinical examinations:** In the previous surveys, we reached consensus (96% agreed) **that the definition should include a list of relevant important clinical signs/changes in examinations of which a patient should have a combination** but NOT ALL, to be considered as having a sinonasal exacerbation. We also reached consensus (more than 80% agreed or strongly agreed) that the following 2 clinical signs/changes in examinations should be included as an indication of a sinonasal exacerbation**: i) mucopurulent nasal discharge or ii) increased mucus production or postnasal drip.**

**Decision to treat and resolution of changes:** In the last survey we reached consensus (more than 80% agreed) that **decision to treat not necessarily with antibiotics but also with increased upper airway clearance or other medication, could be included in the definition as a less important point** (e.g. possibly a minor criterion). Regarding the resolution of changes, we reached consensus **that an important improvement in symptoms/clinical findings after a period of time should be included in the definition of a sinonasal exacerbation as a less important point** (e.g. possibly a minor criterion). We also reached consensus that **absence of improvement in symptoms/clinical findings does not eliminate the diagnosis of acute sinonasal exacerbation** and we will add this as a clarification to the definition. Based on some comments, the wording was not clear enough to be useful in a clinical trial and the period of time should be further defined.

1. Using the major and minor criteria approach, we will include some criteria which are most important for the definition (major) and some which are less important (minor). A combination of major or minor criteria is needed to reach the definition of sinonasal exacerbation.

**Our suggestion is to include all criteria where we already reached a consensus as major unless we already reached consensus to include them as minor.** Please state if you agree using the following criteria as major or minor. (Agree, include as major/ Disagree, include as minor)

- Reported increase in nasal discharge or change in discharge colour (Agree, include as major/ Disagree, include as minor)
- Reported pain/sensitivity in the sinus region (i.e. around the nose, eyes, on the cheeks or forehead) (Agree, include as major/ Disagree, include as minor)
- Reported blocked nose (Agree, include as major/ Disagree, include as minor)
- Mucopurulent nasal discharge at examination (Agree, include as major/ Disagree, include as minor)
- Increased mucus production or postnasal drip at examination (Agree, include as major/ Disagree, include as minor)
- Decision to treat, not necessarily with antibiotics but also with increased upper airway clearance or other medication (Agree, include as minor/ Disagree, do not include)
- Important improvement in symptoms reported by the patient or parent or in clinical findings in case further examination is possible, after a period of 14 days (Agree, include as minor/ Disagree, do not include)

2. Would you like to include any of the following criteria (symptoms or signs/changes in examination), which reached more than 50% but less than 80% consensus or were suggested to discuss again, as minor criteria in the definition?

Please state from the list below which you think should be minor criterion and which should not be included (Agree, include as minor/ Disagree, do not include)

- Reduced quality of life evaluated by any sinonasal specific quality of life questionnaire
- Reported decreased sense of smell
- Signs of acute complication (e.g. orbital infection/abscess, meningitis, cerebral infection, cranial nerve palsy) at examination
- Acute frontonasal / maxillary tenderness at examination

3. We previously listed 5 potential major criteria and 6 potential minor. **Our suggestion is that the definition for sinonasal exacerbation should include a) 3 major or b) 2 major and at least 2 minor criteria.**

Do you agree? Yes/ No

4. If you do not agree, please explain your opinion and make an alternative suggestion (text)
5. Do you have any further comments for the sinonasal definition? (text)
XIII. **Otologic exacerbation in people with PCD (linked to the ears)**

In the third survey, we reached consensus (95% agreed) that exacerbations from the ears are less common than chronic problems in adults with PCD, however when they occur, they can be an important problem.

**Change in symptoms:** In the previous surveys, we reached consensus (100% agreed) that **the definition should include a list of relevant important symptoms of which a patient should have a combination** but NOT ALL, to be considered as having an otologic exacerbation. We also reached consensus (more than 80% agreed or strongly agreed) that the following 3 symptoms (or worsening in them) should be included as an indication of an otologic exacerbation: i) **ear sensitivity/pain, ii) ear discharge or iii) hearing problems.**

**Changes in clinical examinations:** In the previous surveys, we reached consensus (92% agreed) that **the definition should include a list of relevant important clinical signs/changes in examinations of which a patient should have a combination** but NOT ALL, to be considered as having an otologic exacerbation. We also reached consensus (more than 80% agreed or strongly agreed) that the following 2 clinical signs/changes in examinations be included as an indication of an otologic exacerbation: i) **ear discharge or ii) signs of otitis media in otoscopy** (i.e. erythema, collection).

**Decision to treat and resolution of changes:** In the last survey we reached consensus (more than 80% agreed) that decision to treat not necessarily with antibiotics but also with other medication, could be included in the definition as a less important point (e.g. possibly a minor criterion). Regarding the resolution of changes, **we did not reach consensus that an important improvement in symptoms/clinical findings after a period of time should be included in the definition** as a less important point (e.g. possibly a minor criterion), **as it is not always possible** (e.g. in case of a perforated eardrum). However 77% of participants agreed with the sentence above. **We reached consensus that absence of improvement in symptoms/clinical findings does not eliminate the diagnosis of acute otologic exacerbation** and we will add this as a clarification to the definition. Based on some comments, the wording was not clear enough to be useful in a clinical trial and the period of time should be further defined.

6. Using the major and minor criteria approach, we will include some criteria which are most important for the definition (major) and some which are less important (minor). A combination of major or minor criteria is needed to reach the definition of otologic exacerbation.

**Our suggestion is to include all criteria where we already reached a consensus as major unless we already reached consensus to include them as minor.** Please state if you agree using the following criteria as major or minor (Agree, include as major/ Disagree, include as minor)

- Reported ear sensitivity/pain (Agree, include as major/ Disagree, include as minor)
- Reported ear discharge (Agree, include as major/ Disagree, include as minor)
- Reported hearing problems (Agree, include as major/ Disagree, include as minor)
- Ear discharge at examination (Agree, include as major/ Disagree, include as minor)
- Signs of otitis media in otoscopy (i.e. erythema, collection) (Agree, include as major/ Disagree, include as minor)
- Decision to treat, not necessarily with antibiotics but also with other medication (Agree, include as minor/ Disagree, do not include)

7. Would you like to include any of the following criteria, which reached more than 50% but less than 80% consensus or were suggested to discuss again, as minor criteria in the definition? Please state from the list below which you think should be minor criterion and which should not be included. (Agree, include as minor/ Disagree, do not include)

- Reported feeling of fullness in the ears
- Signs of acute complication (mastoiditis, meningitis, cerebral abscess, facial or other cranial nerve palsy) at examination
- Impaired hearing tested by pure-tone audiometry
- Perforated eardrum at examination
- Horizontal nystagmus at examination
- Important improvement in symptoms reported by the patient or parent or in clinical findings in case further examination is possible, after a period of 14 days

8. We previously listed 5 potential major criteria and 7 potential minor. **Our suggestion is that the definition for otologic exacerbation should include a) 3 major or b) 2 major and at least 2 minor criteria.**

Do you agree? Yes/ No

9. If you do not agree, please explain your opinion and make an alternative suggestion (text)
10. Do you have any further comments for the otologic definition? (text)
XIV. **These last points relate to BOTH definitions (sinonasal and otologic):**

In the last round of the survey we reached consensus (85% agreed) that all clinical signs/changes to clinical examinations should be assessed in relation to previous examinations. This was highlighted as particularly important for research settings and will be added as a clarification to the definitions.

11. We also received comments highlighting that realistically clinical practice may differ substantially than research practices, especially for patients residing far from the ENT specialist who follows them for PCD. For this reason, we suggest to include the following clarification for both definitions:

“These definitions are aimed to be used in research settings, especially in clinical trials, to define a sinonasal or otologic exacerbation in patients with PCD”.

We will also add a section in the manuscript discussion, explaining that the definitions could also be useful in clinical practice, discussing specific points that might differ, such as a different number of major or minor criteria.

Do you agree? Yes/ No

12. If you do not agree, please explain and suggest a different wording (text)
13. Would you like us to organise a zoom call to discuss further the final definitions? Yes/No
14. Is there anything else you would like to add in this last survey? (text)

**Thank you for completing this survey and for participating in this project! Please contact** **if you have any questions.**
