## Supplemental Table for "Definition of sinonasal and otologic exacerbation in patients with primary ciliary dyskinesia - an expert consensus"

**Table S1:** Composition of the consensus panel and the patient volunteer group and participation to the electronic Delphi surveys

| Members of consensus panel (N=24) | | Responded to Survey 1  (N=24) | Responded to Survey 2  (N=23) | Responded to Survey 3  (N=23) | Responded to Survey 4  (N=21) |
| --- | --- | --- | --- | --- | --- |
| ENT specialist | 17 | 17  1  1  2  1  1  2  100% | 16  1  1  2  1  1  2  96% | 16  1  1  2  1  1  2  96% | 15  0  1  2  1  1  2  88% |
| Paediatric pulmonologist | 1 |  |  |  |  |
| Adult pulmonologist | 1 |  |  |  |  |
| Physiotherapist | 2 |  |  |  |  |
| Clinical epidemiologist | 1 |  |  |  |  |
| Nurse specialist | 1 |  |  |  |  |
| Patient representatives | 2 |  |  |  |  |
| Patient volunteers | - | 5 | 3 | 3 | 2 |

ENT: Ear-Nose-Throat

Patient volunteers were not members of the consensus panel; they were encouraged to participate to the surveys but there was no required minimum number to proceed to the next survey.

**Table S2:** Responses to key questions of eDelphi survey 1

|  | **Strongly disagree** | **Disagree** | **Neither disagree nor agree** | **Agree** | **Strongly agree** | | **Mean** | **% of agreement** |
| --- | --- | --- | --- | --- | --- | --- | --- | --- |
| 1) **Sinonasal exacerbation definition** | | | | | | | | |
| **Which of the following components can provide the best definition for an exacerbation from the nose and sinuses:** |  | | | | | | | |
| New symptoms or worsening of baseline symptoms from the nose and sinuses, in isolation | 2 | 3 | 7 | 10 | 7 | | 3.59 | 59 |
| Clinical signs or changes in clinical examinations of the nose and sinuses (e.g. nasal endoscopy), in isolation | 3 | 6 | 7 | 9 | 5 | | 3.34 | 48 |
| Changes in imaging (CT or other) of the nose and sinuses in isolation | 5 | 5 | 9 | 7 | 3 | | 2.93 | 34 |
| Decision of ENT specialist to treat, in isolation | 3 | 7 | 11 | 6 | 1 | | 2.72 | 24 |
| Combination of symptoms and changes in clinical examination | 0 | 0 | 2 | 14 | 13 | | 4.66 | 93 |
| Combination of symptoms and decision of ENT specialist to treat | 1 | 1 | 10 | 11 | 7 | | 3.86 | 62 |
| Combination of symptoms and changes in imaging | 3 | 2 | 5 | 15 | 4 | | 3.52 | 66 |
| Combination of changes in clinical examinations and decision of ENT specialist to treat | 1 | 1 | 14 | 8 | 5 | | 3.24 | 45 |
| Combination of changes in clinical examinations and changes in imaging | 2 | 3 | 10 | 11 | 3 | | 3.45 | 48 |
| Changes in imaging and decision of ENT specialist to treat | 2 | 7 | 12 | 5 | 3 | | 3.00 | 28 |
| Combination of symptoms and changes in clinical examinations and decision of ENT specialist to treat | 0 | 1 | 6 | 5 | 17 | | 4.31 | 76 |
| Combination of symptoms and changes in clinical examinations and imaging | 2 | 1 | 5 | 12 | 9 | | 3.86 | 72 |
| Combination of symptoms and changes in imaging and decision of ENT specialist to treat | 2 | 2 | 10 | 10 | 5 | | 3.48 | 52 |
| Combination of symptoms, changes in clinical examinations and imaging and decision of ENT specialist to treat | 2 | 2 | 6 | 5 | 14 | | 3.93 | 66 |
| **Complete resolution of any changes or return to baseline is an important element for the definition of exacerbations from the nose and sinuses in PCD** | 0 | 2 | 7 | 13 | | 7 | 3.8 6 | 69 |
| 2) **Otologic exacerbation definition** | | | | | | | | |
| **Which of the following components can provide the best definition for an exacerbation from the ears:** |  | | | | | | | |
| New symptoms or worsening of baseline symptoms from the ears, in isolation | 1 | 2 | 8 | 12 | | 5 | 3.52 | 59 |
| Clinical signs or changes in clinical examinations of the ears (e.g. otoscopy, audiometry), in isolation | 1 | 5 | 6 | 12 | | 5 | 3.64 | 59 |
| Decision of ENT specialist to treat, in isolation | 3 | 7 | 12 | 4 | | 3 | 3.17 | 24 |
| Combination of symptoms and changes in clinical examination | 0 | 0 | 1 | 16 | | 12 | 4.38 | 97 |
| Combination of symptoms and decision of ENT specialist to treat | 1 | 1 | 10 | 11 | | 6 | 3.69 | 62 |
| Combination of changes in clinical examinations and decision of ENT specialist to treat | 1 | 1 | 13 | 9 | | 5 | 3.55 | 48 |
| Combination of symptoms and changes in clinical examinations and decision of ENT specialist to treat | 1 | 1 | 3 | 9 | | 16 | 4.41 | 86 |
| **Complete resolution of any changes or return to baseline is an important element for the definition of exacerbations from the ears in PCD** | 1 | 2 | 4 | 19 | | 3 | 3.72 | 76 |

ENT: Ear-Nose-Throat, CT: computed tomography, PCD: primary ciliary dyskinesia

**Table S3:** Responses to key questions of eDelphi survey 2 regarding clinical symptoms

|  | **Strongly disagree** | **Disagree** | **Neither disagree nor agree** | **Agree** | **Strongly agree** | **Mean** | **% of agreement** |
| --- | --- | --- | --- | --- | --- | --- | --- |
| 1) **Should changes in the following symptoms be included as an indication of a sinonasal exacerbation?** | | | | | | | |
| Fever/Temperature above 38°C | 2 | 6 | 11 | 4 | 3 | 3.31 | 27 |
| Blocked nose | 0 | 1 | 1 | 14 | 10 | 4.27 | 92 |
| Increase in nasal discharge or change in discharge colour | 0 | 0 | 0 | 8 | 18 | 4.69 | 100 |
| Decreased sense of smell | 0 | 2 | 6 | 12 | 6 | 3.85 | 69 |
| Pain or sensitivity in the sinus region (i.e. around the nose, eyes, on the cheeks or forehead) | 0 | 2 | 2 | 8 | 14 | 4.31 | 85 |
| Headache | 0 | 2 | 9 | 13 | 2 | 3.58 | 58 |
| Malaise/fatigue | 1 | 6 | 9 | 6 | 4 | 3.23 | 38 |
| Increased cough | 0 | 6 | 6 | 11 | 3 | 3.42 | 54 |
| Sore throat | 5 | 6 | 9 | 3 | 1 | 2.54 | 15 |
| Bad breath | 1 | 8 | 8 | 6 | 2 | 3.00 | 31 |
| Bad sleep | 2 | 5 | 5 | 13 | 1 | 3.23 | 54 |
| Reduced QoL evaluated by any sinonasal specific QoL questionnaire | 1 | 0 | 6 | 10 | 9 | 4.00 | 73 |
| 2) **Should changes in the following symptoms be included as an indication of an otologic exacerbation?** | | | | | | | |
| Fever/Temperature above 38°C | 3 | 3 | 10 | 8 | 2 | 3.12 | 38 |
| Ear sensitivity or pain | 0 | 1 | 1 | 13 | 11 | 4.31 | 92 |
| Ear discharge | 0 | 1 | 1 | 9 | 15 | 4.46 | 92 |
| Tinnitus or ringing in the ears | 1 | 4 | 12 | 9 | 0 | 3.12 | 35 |
| A feeling of fullness in the ears | 1 | 2 | 3 | 15 | 5 | 3.81 | 77 |
| Hearing problems | 1 | 0 | 3 | 14 | 8 | 4.08 | 85 |
| Difficulty in communication | 0 | 5 | 12 | 6 | 3 | 3.27 | 35 |
| Malaise/fatigue | 2 | 6 | 12 | 5 | 1 | 2.88 | 23 |
| Headache | 2 | 7 | 10 | 5 | 2 | 2.92 | 27 |
| Dizziness/vertigo | 0 | 3 | 12 | 8 | 3 | 3.42 | 42 |

QoL: quality of life. Symptoms in red were ranked as the most important in their category.

**Table S4:** Responses to key questions of eDelphi survey 2 regarding clinical signs and examinations

|  | **% of agreement** |
| --- | --- |
| 1) **Should the following clinical signs/changes in examinations be included as an indication of a sinonasal exacerbation?** |  |
| Erythema or oedema of nasal mucosa | 54 |
| Mucopurulent nasal discharge | 100 |
| Increased mucus production or postnasal drip | 92 |
| Swelling of inferior turbinates | 42 |
| Appearance or swelling of nasal polyps | 58 |
| Positive microbiology culture from nasal or sinus discharge | 35 |
| Reduced sense of smell measured by any olfactory test | 27 |
| Signs of acute complication (e.g. orbital infection or abscess, meningitis, cerebral infection, cranial nerve palsy) | 72 |
| Negative allergy tests (to exclude allergic causes) | 27 |
| Blood tests for indications of infection (e.g. neutrophils) | 38 |
| 2) **Should the following clinical signs/changes in examinations be included as an indication of an otologic exacerbation?** |  |
| Perforated eardrum | 62 |
| Ear discharge | 92 |
| Nystagmus | 35 |
| Signs of otitis media in otoscopy (i.e. erythema, collection) | 92 |
| Pathologic tympanometry findings | 50 |
| Signs of acute complication (mastoiditis, meningitis, cerebral abscess, facial or other cranial nerve palsy) | 69 |
| Impaired hearing tested by pure-tone audiometry (air and bone-conduction) | 69 |
| Positive microbiology culture from ear discharge | 46 |
| Blood tests for indications of infection (e.g. neutrophils) | 23 |
| Clinical signs/examinations in red were ranked as the most important in their category. |  |

**Table S5:** Responses to key questions of eDelphi survey 3 regarding highly voted items that have not reached consensus on previous rounds

|  | **% of agreement** |
| --- | --- |
| 1) **Should changes in the following symptoms be included as an indication of a sinonasal exacerbation?** |  |
| Reduced QoL evaluated by any sinonasal specific QoL questionnaire | 50 |
| Decreased sense of smell | 58 |
| Headache | 42 |
| Bad sleep | 15 |
| Cough | 15 |
| 2) **Should the following clinical signs/changes in examinations be included as an indication of a sinonasal exacerbation?** |  |
| Erythema or oedema of nasal mucosa | 46 |
| Appearance or swelling of nasal polyps | 46 |
| Signs of acute complication (e.g. orbital infection or abscess, meningitis, cerebral infection, cranial nerve palsy) | 52 |
| 3) **Should changes in the following symptoms be included as an indication of an otologic exacerbation?** |  |
| A feeling of fullness in the ears | 58 |
| 4) **Should the following clinical signs/changes in examinations be included as an indication of an otologic exacerbation?** |  |
| Perforated eardrum | 54 |
| Pathologic tympanometry findings | 38 |
| Signs of acute complication (mastoiditis, meningitis, cerebral abscess, facial or other cranial nerve palsy) | 46 |
| Impaired hearing tested by pure-tone audiometry (air and bone-conduction) | 62 |
| Pathologic pneumatic otoscopy | 35 |
| QoL: quality of life. |  |
